# Malaria temporal dynamic clustering for surveillance and intervention planning

**DOI:** 10.1101/2023.03.24.23287690

**Authors:** Eva Legendre, Laurent Lehot, Sokhna Dieng, Stanislas Rebaudet, Aung Myint Thu, Jade D Rae, Gilles Delmas, Florian Girond, Vincent Herbreteau, François Nosten, Jordi Landier, Jean Gaudart

**Affiliations:** Aix Marseille Univ, IRD, INSERM, SESSTIM, Aix Marseille Institute of Public Health, ISSPAM, Marseille, France; Hôpital Européen Marseille, Marseille, France; Shoklo Malaria Research Unit, Mahidol Oxford Tropical Medicine Research Unit, Mae Sot, Thailand; Mahidol-Oxford Tropical Medicine Research Unit, Faculty of Tropical Medicine, Mahidol University, Bangkok, Thailand; Centre for Tropical Medicine and Global Health, Nuffield Department of Medicine Research building, University of Oxford Old Road campus, Oxford, UK; Institut de Recherche pour le Développement, UMR 228 Espace-Dev (IRD, UA, UG, UM, UR), Phnom Penh, Cambodia; Institut Pasteur du Cambodge, Phnom Penh, Cambodia; La Timone Hospital, BioSTIC, Biostatistics and ICT, Marseille France

**Keywords:** Seasonal malaria, temporal dynamics, clustering

## Abstract

**Background:** Targeting interventions where most needed and effective is crucial for public health. Malaria control and elimination strategies increasingly rely on stratification to guide surveillance, to allocate vector control campaigns, and to prioritize access to community-based early diagnosis and treatment (EDT). We developed an original approach of dynamic clustering to improve local discrimination between heterogeneous malaria transmission settings.

**Methods:** We analysed weekly malaria incidence records obtained from community-based EDT (malaria posts) in Karen/Kayin state, Myanmar. We smoothed longitudinal incidence series over multiple seasons using functional transformation. We regrouped village incidence series into clusters using a dynamic time warping clustering and compared them to the standard, 5-category annual incidence standard stratification.

**Results:** We included 1,115 villages from 2016 to 2020. We identified eleven *P. falciparum* and *P. vivax* incidence clusters which differed by amplitude, trends and seasonality. Specifically the 124 villages classified as “high transmission area” in the standard *P. falciparum* stratification belonged to the 11 distinct groups when accounting to inter-annual trends and intra-annual variations. Likewise for *P. vivax*, 399 “high transmission” villages actually corresponded to the 11 distinct dynamics.

**Conclusion:** Our temporal dynamic clustering methodology is easy to implement and extracts more information than standard malaria stratification. Our method exploits longitudinal surveillance data to distinguish local dynamics, such as increasing inter-annual trends or seasonal differences, providing key information for decision-making. It is relevant to malaria strategies in other settings and to other diseases, especially when many countries deploy health information systems and collect increasing amounts of health outcome data.

**Funding:** The Bill & Melinda Gates Foundation, The Global Fund against AIDS, Tuberculosis and Malaria (the Regional Artemisinin Initiative) and the Wellcome Trust funded the METF program.

## Introduction

During malaria control or pre-elimination phases, the World Health Organization (WHO) recommends countries to rely on a stratification process in order to allocate resources and interventions. Stratification aims to produce discrete maps differentiating areas based on transmission intensity and receptivity.^1^ Transmission intensity can be measured directly by entomological parameters (entomological inoculation rate) but require extensive efforts and significant resources.^2,3^ In practice, clinical malaria incidence over one year, also called “annual parasite incidence” (API) is often used as a proxy of transmission. Receptivity is a complex concept without a straightforward quantitative measurement.^3^ In the context of prevention of reintroduction, it mainly refers to the ability of an ecosystem to enable malaria transmission even if it is not ongoing: presence of competent vectors, suitable climate and susceptible population.^1^ Vectorial capacity can provide important insights on receptivity, but due to lack of large scale entomological information, receptivity is often approximated using environmental (meteorological/landscape) data.^1,3^ Depending on data availability, stratification can be performed from broad (region, district) to fine (village, health area) geographical scales to fit operational needs.

In the Greater Mekong Subregion (GMS), malaria persists in some high transmission foci where *Plasmodium falciparum* and *P. vivax* are responsible of most morbidity and mortality.^4^ *P. falciparum* elimination in this area has been prioritized in response to the emergence of artemisinin resistance.^4–6^

Myanmar remains the country with the highest malaria burden in the GMS, especially in mountainous borderlands. In accordance with WHO technical guidelines, Myanmar planned its 2016-2030 elimination strategy relying on API to stratify its regions and townships for operational planning.^7^ Townships are not the finest administrative division in Myanmar; it is subdivided in village tracts and villages. This stratification method is easily implemented based on routine data collections but the stratification may not be sufficient to adapt and optimize surveillance and intervention. When malaria control progresses, malaria transmission becomes increasingly heterogeneous and unstable, for example due to local differences in the environment sustaining transmission or in the impact of interventions. The complex settings of malaria elimination also imply sporadic epidemics, more unexpected temporal dynamics not addressed with classical approaches, and need to assess the whole time series evolution. To consider this variability, it is necessary to develop tools that describe more accurately malaria dynamics over several years.

According to WHO and Myanmar NMCP report in 2015, stratification at township level identifies Karen state mostly as a moderate transmission area with high risk in the northern township.^8^ This state, located at the Myanmar-Thailand border, is a typical GMS elimination setting combining low incidence areas with persisting high transmission hotspots. In this state, the Malaria Elimination Task Force (METF) was launched in 2014. The program offered early diagnostic and treatment in over 1200 malaria posts (MPs) in four townships and targeted mass drug administration (MDA) in selected high prevalence village.^9,10^ It offered the opportunity to describe malaria heterogeneity at a finest geographical scale operationally relevant - the village.

This study proposed a method which allow stratification in settings where multiple, different malaria seasonality coexists, and where epidemiology and dynamics can change due to interventions or environmental drivers. We compared two malaria incidence stratifications (routine standard versus temporal dynamic clustering method) to discriminate malaria transmission settings at village scale. The temporal dynamic clustering method aimed to identify hotspots of transmission and temporal dynamics heterogeneities at village scale over several years and for each malaria species. It will help to further understand malaria epidemiology in the Karen state and optimize surveillance and interventions beyond conventional API-based stratification. This required to analyse hundreds of heterogeneous time series simultaneously without statistical assumptions. We propose a temporal dynamic clustering methodology to overcome this methodological hurdle.

## Method

### Study design and setting

In this study, we analysed longitudinal records of *P. falciparum* and *P. vivax* malaria cases by malaria post (MP) of the Malaria Elimination Task Force (METF), between 2016 and 2020 in the Karen state of Myanmar along the Thailand border. 95% of villages identified in the target region hosted a malaria post, the vast majority opening between May 2014 and August 2016 (n=1057, 95%).^11^ MP systematically tested fever cases using a *P. falciparum-P. vivax* rapid diagnostic test (RDT) and sent weekly reports specifying the number of *P. falciparum* and *P. vivax* cases diagnosed.^9^ At the time of the analysis, malaria records were available until March 2020.

### Data of malaria incidence rate

For each village, we estimated *P. falciparum* and *P. vivax* observed incidence as the weekly or annual case count recorded by MPs divided by the village population during study period. The population was estimated using the number of households in the village and an average of 5 members per household according to Myanmar census data.^12^

### Malaria stratification on annual parasite incidence (API) in 2019

For each village, API was calculated as the sum of malaria cases in a year divided by the number of people at risk in a year and reported as cases per 1000 person.year at risk. Following WHO GMS guidelines, Myanmar national plan for malaria elimination distinguished 5 strata based on API (*P. falciparum* and *P. vivax* cases combined) in 2019: transmission free area (API=0 /1000 individuals under surveillance for more than 3 years), potential transmission area (API=0), low transmission area (API<1), moderate transmission area (API between 1-5) and high transmission area (API>5).^7,13^

### Statistical analysis: the temporal dynamic clustering

In the region, malaria is seasonal with patterns which could vary between areas and time. The objective of the temporal dynamic clustering method is to group malaria dynamics with similar seasonality, amplitude, shape, and by associating malaria outbreaks occurring in the same season. The method includes 3 successive steps: 1/ functional transformation to smooth time series; 2/ dynamic time warping (DTW) metric calculation on functional data; 3/ partitioning around medoid (PAM) clustering on DTW matrix **(Sup. Fig. 1)**.

The development of the temporal dynamic clustering responded to 3 challenges. First, the clustering method needed to consider not only amplitude but also malaria seasonality (i.e. intra-annual variability), curve shape, and allowing time lags to associate outbreaks occurring with few weeks of interval during the same season. Based on our previous comparison of metrics in the context of malaria, DTW metric was the most appropriate.^14^ Second, DTW algorithm performance is sensitive to sharp irregularities.^15^ Malaria time series at village scale were sharp and noisy, and required smoothing. In this complex elimination setting, environmental changes (meteorological, landscape, etc.), socio-political events or METF interventions occurrence induced differential dynamic variations and invalidated the stationarity assumption. It prevented the use of classical time series methods (e.g. ARIMA). Functional data transformation allowed to remove sharp irregularities, with smoothing parameters identical and optimized to the entire set of time series, and without stationarity assumptions.

Analyses were conducted using R 4.0 (R Development Core Team, R Foundation for Statistical Computing, Vienna, Austria) and {fda}, {TSclust} and {dtw} packages. An R script is available in supplementary material to facilitate the implementation of the clustering method (**Sup. Met. 1**).

#### Preliminary settings: village and time frame selection

We grouped cases and population denominators from MPs located less than 500m away from each other in “villages” - the statistical unit of this study.

Simultaneous functional transformation required time series with identical length. Because of MP were set up gradually, we selected a common study period based on calendar dates. We chose the period to include a maximal number of villages having at least one malaria case and to yield the period with the longest possible duration.

We excluded villages with more than two missing successive weekly report (>1% missing data) from the analysis. For villages with less than two consecutive weeks missing, we imputed missing weekly reports by the average of the number of cases from the following and the previous weeks. Missing reports were due to omission, phone network problem, problem of access, loss of the report, etc. We excluded villages without population denominator (missing household count) from the analysis.

#### 1/ Smoothing malaria time series using functional data transformation

Incidence time series were sharp, noisy, and sometimes seasonal. We used functional data to smooth simultaneously all the time series. For each time series, we estimated a function from its observed values. Every y time series of i weeks was transformed by a smooth function x(t), which was a linear combination of elementary function, i.e. cubic B-splines, and an error term **(Equation 1, 2)**. Cubic B-splines functions were grouped in a function basis ϕ_k_ (t) containing k functions. As recommended, the number of functions k was equal to the number of weeks included in the study period.^16^ Coefficients c were estimated by minimizing the penalized (PEN) sum of squared errors (SSE) **(Equation 3)**. The penalty was on the roughness of the function x(t) (i.e. measures the curvature of the curve). The smoothing parameter λ, indicating the emphasis of the penalization, was optimized by minimizing the sum of the GCV criterion of all v villages using a set of values of log(λ) ranging from 0 to 10 with a step of 0.25 **(Equation 4)**.^16^

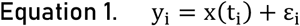

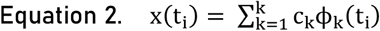

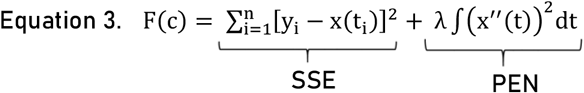

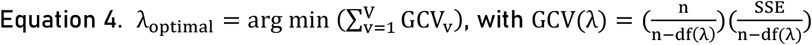

A square root transformation was applied before functional transformation to the incidence rate time series because of large-scale amplitude to the incidence rates.^14^

#### 2/ Distance between functional time series using the DTW metric

The dynamic time warping metric is a flexible metric, which distinguished two time series according to their amplitude and phases by considering time lags. The algorithm calculated distance between two villages after compressing or stretching functional time series locally to make them as similar as possible. This metric can be parametrized to specify the time-window of the stretch. We chose a three months window to associate malaria outbreaks belonging to the same transmission season (cold or rainy season).^17,18^

#### 3/ PAM clustering on DTW matrix

We applied partitioning around medoid (PAM) clustering to the distance matrix calculated with the DTW metric. PAM algorithm is similar to the k-means algorithm, except that the centre of each class, called “medoid”, corresponds to an observed time series and it is compatible with the DTW metric (i.e. a non-Euclidian distance).^19–21^ We named each cluster based on visual assessment of its distinctive characteristics.

Minimization of the intra-class inertia average (I) and parsimony principle determined the optimal number of clusters G. The conditional intra-class inertia (CI) was estimated for each G cluster by the average of the DTW distance between the n villages (v) included in the cluster and its medoids (M) **(Equation 5)**.

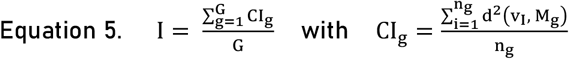

### Sensitivity analysis

Simple approaches to improve API stratification were (i) to divide the “high transmission area” category (>5 cases/1000py) in sub-categories and (ii) to consider stratification results over several years. We compared our temporal dynamic clustering with a third stratification from 2016 to 2019 based on API stratification but subdividing the high transmission area category according to incidence quartiles.

## Results

### API stratification for Myanmar in 2019

Since 2014, 1,205 MPs were set up by the METF program and were grouped in 1,115 villages spreading over 4 townships. In 2019, 1,032 villages had active MP reporting. The median population in the villages was 200 and the IQR was 115-360. For the Northern township where most malaria cases occurred, the median village population was 142 (IQR=95-215).

According to API stratification on both malaria species in 2019, 48.4% (n=499) of the villages corresponded to a high transmission area and 28.9% (n=298) to a transmission-free area. Taking into account only *P. falciparum* cases, the proportion of villages in a transmission-free area increased to 57,1%, twice more than in the overall malaria stratification. On the contrary, the proportion of villages in a high transmission area were divided by more than 3 to reach 14.5%. Considering *P. vivax* only, proportions were similar to the overall malaria stratification. This implied that malaria stratification is driven by *P. vivax* cases, which represented 91,9% of the total malaria burden (**Tab. 1 & Fig. 1**).

**Table 1.** Annual parasite incidence (API, equivalent to annual malaria cases incidence /1000 individuals under surveillance) stratification in 2019 at village level. N(%).

|  | Malaria (both species) | *P. falciparum* | *P. vivax* |
| --- | --- | --- | --- |
| Transmission free area (API=0 for more than 3 years) | 298 (28.9) | 586 (57.1) | 324 (31.4) |
| Potential transmission area (API=0) | 167 (16.2) | 250 (24.4) | 154 (14.9) |
| Low transmission area (API<1) | 3 (0.3) | 1 (0.1) | 3 (0.3) |
| Moderate transmission area (API=1-5) | 65 (6.3) | 40 (3.9) | 64 (6.2) |
| High transmission area (API>5) | 499 (48.4) | 149 (14.5) | 487 (47.2) |

**Figure 1.**
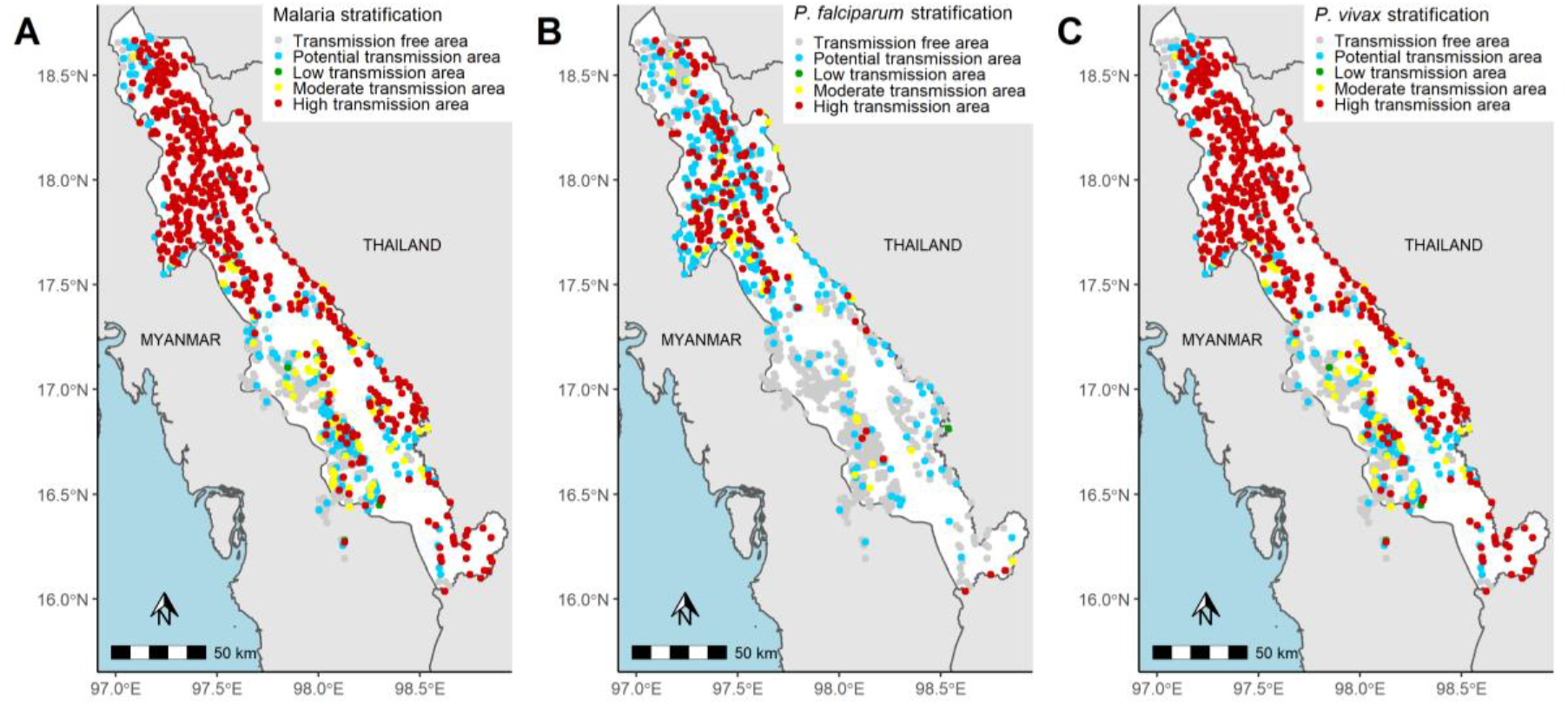
Map of API stratification on annual parasite incidence in 2019 at village level for A) malaria (both species), B) *P. falciparum* and C) *P. vivax*.

### Temporal dynamic clustering

#### Village and study period selection

Seven villages without population denominator were excluded. The period from March 2016 to February 2020 maximized the number of villages included and the time series duration, with 676 villages followed over 206 weeks. Fourteen villages with >2 successive missing reports were excluded. Missing reports occurred rarely over time and villages (118 missing of 136,372 reports expected, i.e. 0.086%). The analysis included 662 villages **(Sup. Fig. 2)**. Of 662 villages, 16.2% (n=107) belonged to the malaria transmission free stratum, 16.3% (n=108) belonged to the potential transmission stratum, 0.1% (n=1) to low, 5.9% (n=39) to moderate and 61.5% (n=407) to high transmission area strata.

#### *P. falciparum* clustering

The 662 *P. falciparum* incidence rate time series were simultaneously transformed into functional data (k=206, λ=100, GCV=196). The temporal dynamic clustering identified 11 clusters (intra-class inertia=104) **(Sup. Fig. 3)**.

Most villages (81%, n=539) belong to the “very low” cluster, with a near zero *P. falciparum* incidence between 2016 and 2020. The “low” cluster regrouped 46 (7%) villages with sporadic cases and a low incidence rate. Two clusters exhibited an incidence peak during the rainy season in 2017 (0.8%, n=5) or 2018 (2%, n=13). 40 villages exhibited a single incidence peak during the cold season and were grouped in three clusters according to the year: 2016 (2.7%, n=18), 2017 (2.6%, n=17) or 2018 (0.8%, n=5). One cluster including 10 villages (1.5%) presented a decreasing trend with peaks in both cold and rainy seasons. Two clusters regrouped villages with a high incidence rate until September 2018 (n=5 or 0.8% and n=3 or 0.5%). The 11^th^ cluster corresponded to a single village (**Fig. 2**).

**Figure 2.**
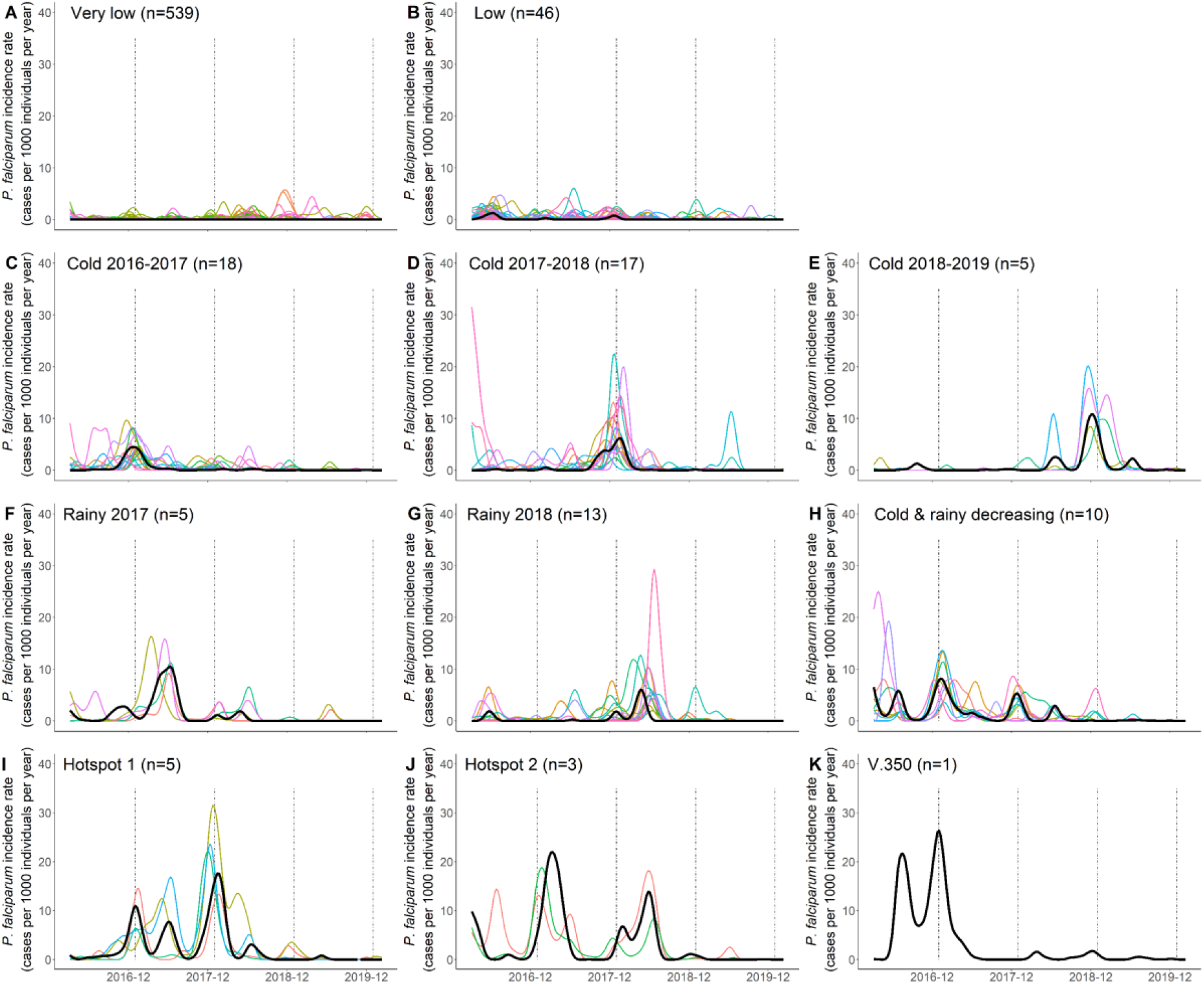
*P. falciparum* clustering. Temporal dynamic clustering identified eleven clusters. Black line corresponds to the central village (medoid) of each cluster. Coloured lines represent functional incidence rate of all villages included in the cluster across study period. When clusters contained only one village, only the medoid (black line) is presented corresponding to the time series of the village.

When compared to API stratification, *P. falciparum* dynamic clustering distinguished villages according to both their current and previous incidence. Within the potential transmission stratum, the clustering distinguished villages with a consistently low incidence (n=22) from villages where incidence recently dropped after persisting at high levels (hotspot, n=2). Likewise, it identified 4 patterns in the high transmission stratum: 54 villages with a past incidence near zero, 36 villages with a past single outbreak, 6 villages with a decreasing tendency and 6 hotspots (**Tab. 2**).

**Table 2.**
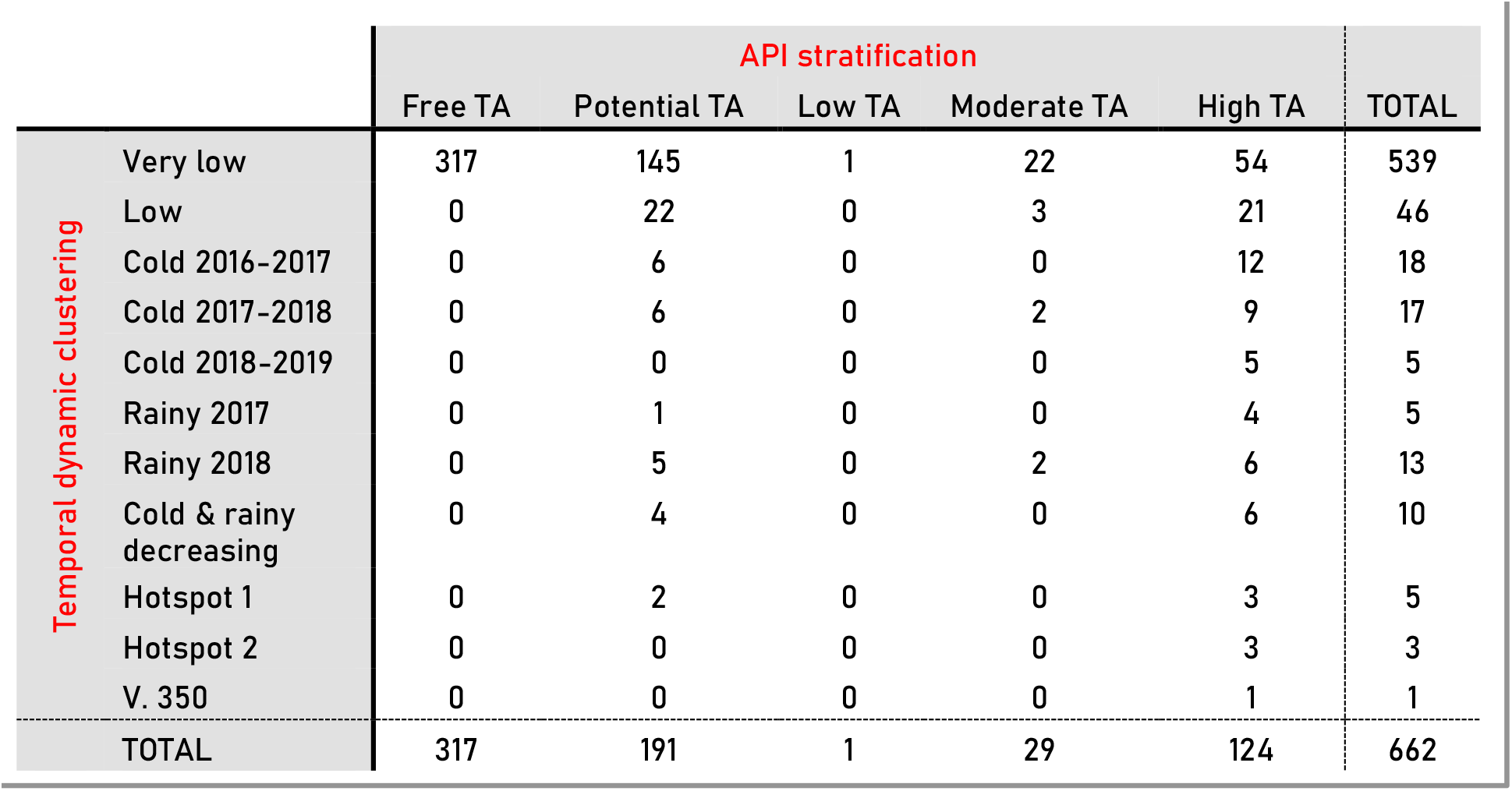
Comparison between WHO stratification and temporal dynamic clustering for *P. falciparum*. The number of villages for each cluster/stratum is presented (TA: transmission area).

#### *P. vivax* clustering

The 662 *P. vivax* incidence rate series were simultaneously transformed into functional data (k=206, λ=562, GCV=487.5). The temporal dynamic clustering identified 11 clusters (I=227) (**Sup. Fig. 3**).

A majority of villages (69%, n=457) belonged to the “very low” incidence cluster across the study period. Two clusters presented a low incidence rate. One presented sporadic cases across years (n=69, 10%), whereas the other displayed an increasing trend (n=45, 7%). Two clusters exhibited a marked seasonality in the rainy season and were distinguished by their trend: one decreased over time (n=13, 2%), whereas the other increased (n=23, 3.5%). A cold-season cluster was also identified (n=31, 5%), with its highest incidence during the 2017-18 cold season. Another cluster showed a low incidence until the end of 2017 and a rapid increase to a higher incidence rate after (n=14, 2%). One cluster identified villages with persistent cases (n=7, 1%). Finally, three clusters isolated individual villages (**Fig. 3**).

**Figure 3.**
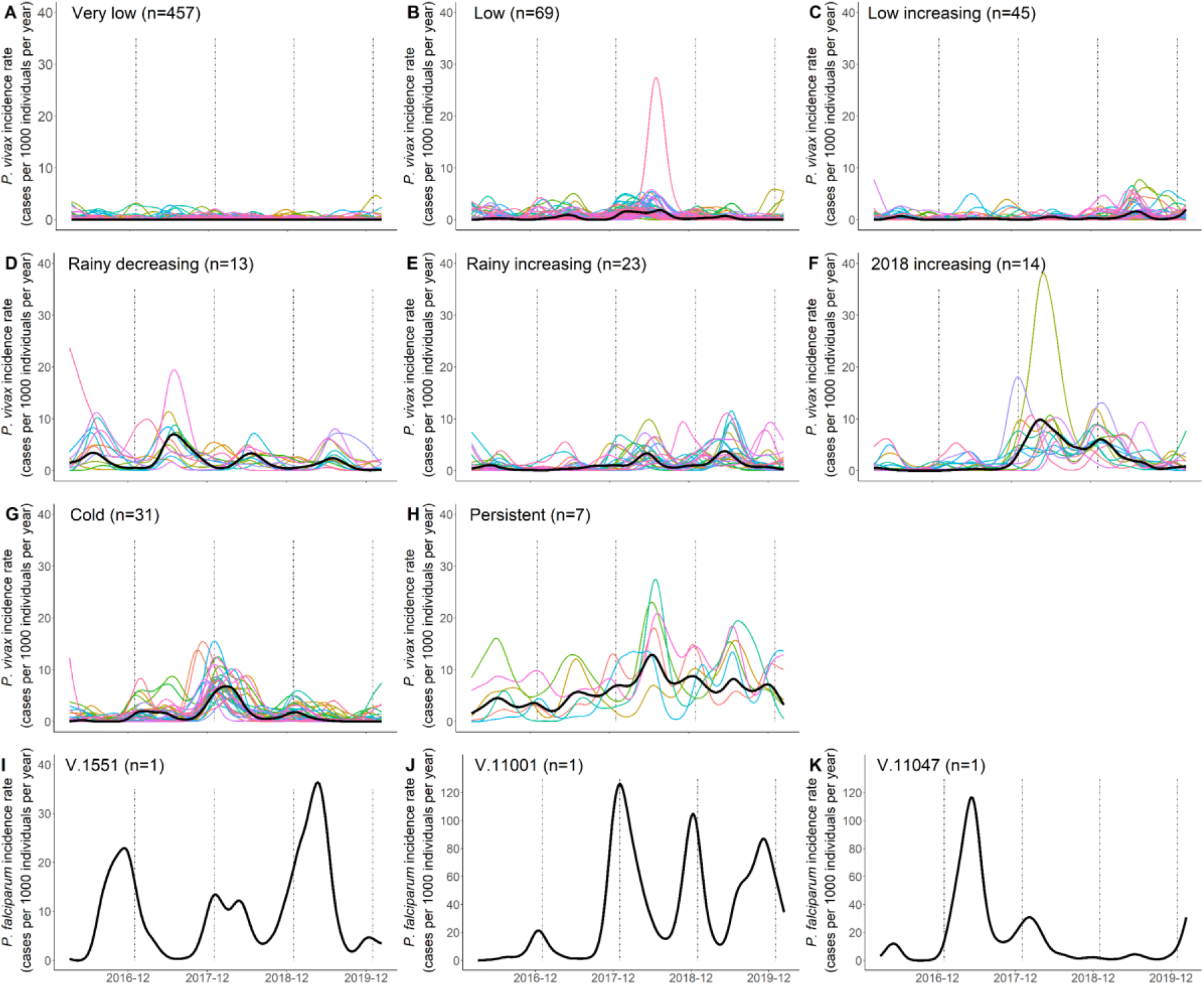
*P. vivax* clustering. Temporal dynamic clustering identified eleven clusters. Black line corresponds to the central village (medoid) of each cluster. Coloured lines represent functional incidence rate of all villages included in the cluster across study period. When clusters contained only one village, only the medoid (black line) is presented corresponding to the time series of the village.

When compared to Myanmar API stratification, *P. vivax* dynamic clustering identified 10 different patterns within the high transmission stratum according to past incidence amplitude, tendency, and seasonality. Conversely, the “very low” cluster included the other 4 strata. (**Tab. 3)**

**Table 3.** Comparison between API stratification and temporal dynamic clustering for *P. vivax*. The number of villages for each cluster/stratum is presented (TA: transmission area).

|  |  | API stratification |  |  |  |  | TOTAL |
| --- | --- | --- | --- | --- | --- | --- | --- |
|  |  | Free TA | Potential TA | Low TA | Moderate TA | High TA |  |
| Temporal dynamic clustering | Very low | 120 | 103 | 1 | 38 | 195 | 457 |
|  | Low | 0 | 1 | 0 | 0 | 68 | 69 |
|  | Low increasing | 0 | 0 | 0 | 0 | 45 | 45 |
|  | Rainy decreasing | 0 | 0 | 0 | 0 | 13 | 13 |
|  | Rainy increasing | 0 | 0 | 0 | 0 | 23 | 23 |
|  | Cold | 0 | 0 | 0 | 0 | 31 | 31 |
|  | 2018 increasing | 0 | 0 | 0 | 0 | 14 | 14 |
|  | Persistent | 0 | 0 | 0 | 0 | 7 | 7 |
|  | V. 1551 | 0 | 0 | 0 | 0 | 1 | 1 |
|  | V. 11001 | 0 | 0 | 0 | 0 | 1 | 1 |
|  | V. 11047 | 0 | 0 | 0 | 0 | 1 | 1 |
|  | TOTAL | 120 | 104 | 1 | 38 | 399 | 662 |

#### Geographical distribution of *P. falciparum* and *P. vivax* clusters

All but one villages which did not belong to the “very low” cluster of *P. falciparum* incidence were grouped in the Northern part of the Karen state with one specific group at the extreme north (in yellow) corresponding to an outbreak in cold season 2018-19 (**Fig. 4A**). *P. falciparum* hotspots villages (in orange and pink) were concentrated in another specific area.

**Figure 4.**
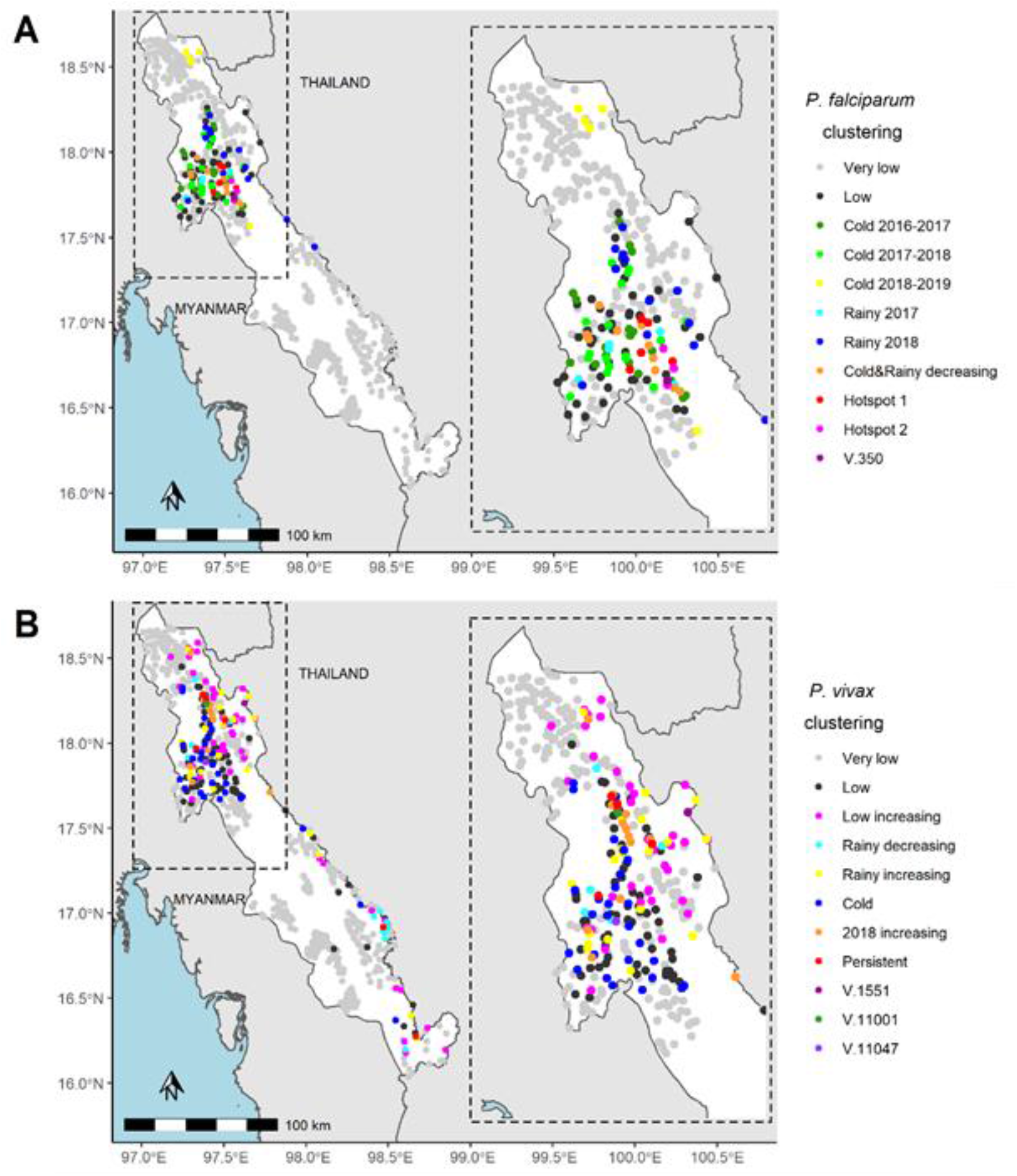
Maps of village in the METF target region according to malaria clustering. A) *P. falciparum* clusters and B) *P. vivax* clusters with a zoom on Northern area.

Concerning *P. vivax*, villages which did not belong to the “very low” cluster of incidence were also grouped in the Northern part and along the Thailand-Myanmar border (**Fig. 4B**). Villages with a *P. vivax* increasing or persistent tendency (in pink, yellow, orange and red) were mostly localized in the Northeast part with a core composed of villages with a brutal incidence increasing in 2018 (in orange) or with persisting high incidence (in red). Another substantial aggregation of villages is present in the Southeast region with one *P. vivax* annual peak only in rainy season and with a decreasing tendency (in turquoise). Temporal dynamic clustering thus led to relevant spatial distribution of clusters.

### Sensitivity analysis

In order to address the wide range of incidence in the high transmission category of the API stratification, we split this category by quartiles: 5-10, 10-30, 30-100, ≥ 100. We compared this 6-category API stratification to the temporal dynamic clustering from 2016 to 2019 **(Sup. Fig. 4-5)**. 6-category stratification distinguished villages according to amplitude, some annual trends and no seasonality. The “Very low” clusters stood out from the others considering API categories (i.e. incidence amplitude). For *P. vivax*, the “2018 increasing” and “Rainy decreasing” clusters were identical considering API categories. For *P. falciparum*, “Cold 2017-2018” and “Rainy 2017” clusters or “Cold 2018-2019” and “Rainy 2018” were also similar.

## Discussion

Based on WHO recommendations, stratification on API (combining *P. falciparum* and *P. vivax*) incidence at village level in 2019 identified 54.7% of the villages in a moderate or high transmission area. According to national guidelines, early diagnostic and treatment should be reinforced with preventive measures in those stratum representing 564 villages.^7^ Analyses distinguishing *Plasmodium* species showed that only 18.4% of the villages still had moderate or high *P. falciparum* transmission. It is consistent with global analysis showing that the proportion of cases attributable to *P. vivax* increased as *P. falciparum* incidence declined.^22^ In co-endemic countries, distinct analysis according to parasite species is necessary to adapt species-specific interventions. Moreover, in a *P. falciparum* pre-elimination setting, combined *P. vivax* survey could inform about *P. falciparum* receptivity (i.e. due to longer persistence) useful to target surveillance and prevent resurgence.

Even by considering *Plasmodium* species, API-based stratification pooled villages belonging to the *P. falciparum* hotspot cluster and low incidence patterns together, and all *P. vivax* dynamic patterns identified in the high transmission area stratum. API-based stratification could lead to coarse targeting interventions because of an insufficient discriminating stratification. By considering incidence past-years evolutions (amplitude level, tendency) and seasonality variations (i.e. intra-annual variability), the temporal dynamic clustering was more discriminant and proved to be more adapted to allocate surveillance and intervention. The clustering allowed to identify villages where additional measures are necessary (e.g. increasing tendency, persisting high incidence, recently reach zero cases), or on the contrary the one where existing interventions appear sufficient (e.g. low incidence decreasing) (**Details in table 4)**.^7^ It also identified intra-annual patterns and distinguished villages with one or two annual peaks, and their seasonality. This could be exploited to adapt supplies and community engagement towards preventive behaviours. It is also of interest to identify transmission bottlenecks and deployed sustainable elimination strategies by highlighting low transmission period.^23^ The temporal dynamic clustering provided rich information about malaria dynamics, which could not be obtained through a simple, or a more complex stratification on API. Sensitivity analyses were conducted by dividing the high transmission stratum in subcategories and considering all API values over the 2016-2020 periods, without equalling the precision of the DTW metric (i.e. seasonality with time lag) (**Sup. Fig. 4, 5**)

**Table 4.** Surveillance and intervention recommendations according to API-based stratification (AS) and temporal dynamic clustering (TDC).

| Stratification | Malaria epidemiology | Surveillance and intervention recommendations |
| --- | --- | --- |
| AS & TDC | Zero cases for few years | Passive surveillance |
| AS & TDC | Recently reach zero cases | Active surveillance (e.g. mobile team) |
| TDC | Low incidence with a decreasing tendency | No additional measures than existing ones |
| TDC | Low incidence with an increasing tendency | Additional interventions (e.g. reinforce preventive measure, early diagnostic & treatment) |
| TDC | Persisting high incidence | Additional interventions (e.g. reinforce preventive measure, early diagnostic & treatment, mass intervention) |
| TDC | Single village with specific dynamics & high incidence | Investigation |

This study presented several strengths. First, the unique dataset collected by the METF community-based MP network allowed studying malaria incidence at an unprecedented granularity (village-scale) in the GMS. Beyond identifying the northern area of the region with the highest malaria burden (already shown elsewhere^8,10^), clustering highlighted localized groups of few villages relevant for operation planning. *P. falciparum* and *P. vivax* dynamics also could be studied separately thanks to data richness. Second, we adapted existing methods into a straightforward, step-by-step analysis workflow allowing the exploration of the temporal dynamics of a large number of time series simultaneously without assuming stationarity. An open-access script makes methodology implementation simple and repeatable in other contexts (**Sup. Met. 1**).

This study presented several limits. Functional transformation smoothing was necessary to transform series into continuous function and to optimize DTW algorithm.^15^ Because MP opened gradually, data was excluded to obtain a set of observations over the same calendar period. In total 395 villages with late MP opening were excluded. However, these villages had in majority extremely low malaria incidence (4.3% villages with >5 *P. falciparum* case/year and 12.7% with >5 *P. vivax* case/year). Likewise, the first months of MP activity were excluded for MP opening earlier than the date chosen to initiate the study. This represented 12 months in average and may have prevented from identifying *P. falciparum* incidence patterns where it decreased quickly after providing access to diagnosis and treatment.

Second, clustering analysed only clinical malaria incidence and thus transmission characteristics. According to WHO guidelines, taking into account malaria receptivity will improve this clustering. Combining *P. falciparum* and *P. vivax* analysis brings some information about receptivity, but this should to be completed with more detailed data including environmental characteristics.

Finally, smaller towns could increase the variance of the incidence. It is a general concern with incidence estimation. Nevertheless, PAM clustering discriminated single outliers with a unique dynamic (e.g. Fig 3. I-K). It highlights villages where specific investigation is necessary (to understand if the unique profile identified corresponds to a truly specific situation or to correct potential data errors).”

The method proposed here provides a thorough tool to exploit longitudinal routine case data and extract an untapped wealth of information for operational planning, allowing to gauge trends, seasonality shifts or periodic outbreaks. We applied it to a fine-grained village-level malaria incidence dataset. This approach relies on village-level data yet unavailable in many settings. Our method could be used to identify longitudinal incidence patterns at coarser scale in a country-wide approach (e.g. village tract or health area). WHO “high burden to high impact” approach promotes targeting areas where malaria burden is highest, leading to stratification beyond district level, e.g. health areas or health facility catchment areas.^24,25^ Other infectious diseases with highly heterogeneous dynamics driven by multiple factors could also be analysed in this workflow.

In conclusion, to be efficient, control and intervention planning need to take into account fine description of malaria dynamics. More sophisticated stratification than API over one year is needed to characterize it. The temporal dynamic clustering is an adapted methodology to support malaria surveillance and intervention planning which could be applied to a large number of spatial units at different scales (health area, health district or region levels) in countries of the GMS, and in other regions of the world, particularly where malaria transmission is highly seasonal (Sahel, Southern Africa…).

## Data Availability

The data analysed for this study are available upon request to the Mahidol Oxford Tropical Medicine Research Unit data access committee: https://www.tropmedres.ac/units/moru-bangkok/bioethics-engagement/data-sharing.

## Acknowledgments

The authors thank the people of Karen/Kayin State whose engagement and participation to the programme was a key element of success. We acknowledge the hard work, dedication and continuous support of all staff, collaborators and colleagues who made this project possible and contributed to implementing it up to the most remote communities. SMRU is part of the Mahidol Oxford University Research Unit, supported by the Wellcome Trust of Great Britain.

## Contributors

JL and JG designed the study. EL, JL and JG designed the analysis plan. EL developed the statistical analysis framework and conducted the analysis, under the supervision of JG, and with methodological contributions of SD and LL concerning functional transformation. AT, JR, JL, FN, GD supervised METF program. AT, JR, JL collected data. FN, VH, FG, GD, JR, AT, JG, SR and JL helped to the interpretation of the findings. EL wrote the first draft of the manuscript with contributions of JL, JG and SR. All authors contributed to review the analysis, wrote the manuscript and approved the final manuscript.

## Availability of data and materials

An open-accessed R script is available to facilitate methodological implementation of the temporal dynamic clustering (**Sup. Met. 1**).

## Abbreviation

CI: conditional intra-class inertia
DTW: dynamic time warping
GMS: Greater Mekong Subregion
I: intra-class inertia
MDA: mass drug administration
METF: Malaria Elimination Task Force
MP: malaria post
NMCP: National Malaria Control Program
PAM: Partitioning around medoids
TA: Transmission area
WHO: World Health Organization

